# What the analysis of causes of death in France in 2020 reveals about the impact of the Covid-19 epidemic

**DOI:** 10.1101/2023.03.07.23286673

**Authors:** Laurent Toubiana, Laurent Mucchielli, Jacques Bouaud, Pierre Chaillot

**Affiliations:** Inserm, Sorbonne Université, Université Sorbonne Paris Nord, UMR S_1142, LIMICS, Paris, France; IRSAN, Institut pour la valorisation des Données de Santé, Paris, France; CNRS, Centre méditerranéen de sociologie, de science politique et d’histoire, UMR 7305, LAMES, Aix-Marseille Universités, France; INSEE, Institut national de la statistique et des études économiques, Paris, France; AP-HP, Assistance Publique-Hôpitaux de Paris, DRCI, Paris, France

**Keywords:** Covid-19, epidemic, pandemic, statistics, causes of death, ICD-10, mortality, excess mortality, cause of death, aging, population structure

## Abstract

**Context:** In 2020, the French population lived under the threat of the Covid-19 epidemic, which would allegedly cause an exceptional excess mortality rate. Mortality data by cause of death for the year 2020 are now available. These data allow us to quantify and qualify the impact of the epidemic. This analysis presents the evolution of the main mortality indicators by cause and by age group. It is intended to demonstrate how 2020 was an exceptional year.

**Materials and methods:** In France, causes of death are labeled according to the International Classification of Diseases (ICD-10) based on medical death certificates. The study focuses on aggregated data for the year 2020 as well as complete data from 1979 to 2017 available online. To estimate excess mortality by cause of death, mortality data were standardized to 2020 to consider changes in the age structure of the population.

**Results:** The year 2020 is marked, on the one hand, by the introduction of “Covid-19” as a cause of death, which accounts for 10.4% of deaths; and, on the other hand, by a strong downward trend in most other causes of death.

**Discussion:** In 2020, we show that the overall number of additional deaths due to the Covid-19 epidemic is paradoxically lower than the number of deaths caused by this disease. According to official sources, the number of Covid-19-labeled deaths is nearly 50% higher than the number of additional deaths. Besides excess mortality in 2020 being modest compared to other years that saw health events and it affecting only individuals over 65 years of age, what could explain that this disease has caused more deaths than additional deaths? This analysis shows that the emergency implementation of the “Covid-19” classification led to many biases. A significant number of deaths, usually labeled for other major causes (e.g., neoplasm, circulatory system diseases) were, in a way, transferred to this new label. This analysis of mortality by cause of death provides quantitative answers to the overestimation of the impact of the Covid-19 epidemic in France.

## Introduction

In early 2020, the first cases of a disease affecting the respiratory tract were detected in France. Some epidemiologists, based on computer models, estimated at the time that the disease, named Covid-19, could cause 500,000 additional deaths in France^1,2^. These estimates were one of the main arguments for the implementation of far-reaching health measures. Lockdowns^3^, curfews, tests, masks, social distancing and, from 27 December 2020, the beginning of a mass vaccination campaign. In this regard, the 2020 Covid-19 epidemic is unprecedented.

The study of mortality data is used to assess the impact of a health event on health. While populations went through 2020 under the threat of an exceptional excess mortality, studies have shown that the excess mortality in 2020 was relatively small^4^. Furthermore, no excess mortality was detected for most of the population (i.e., nearly 80%).

The national statistics on causes of death published by the C*entre d’épidémiologie sur les causes médicales de Décès* (CépiDc) of the *Institut national de la santé et de la recherche médicale* (Inserm) for the year 2020 are now available. The 2020 data on causes of death, especially since the introduction of “Covid-19” as a specific cause, make it possible to quantify and qualify the impact of the epidemic on mortality.

The purpose of our study is to present the evolution of the main mortality indicators by cause and by age group. We will show how 2020 is an exceptional year. We have studied the hierarchy of causes of death in France, as well as the stability of their trends over the past 15 years. We will also show how the analysis of causes of death by age group confirms the analysis of the data published by *L’Institut national de la statistique et des études économiques* (Insee) from the beginning of 2021^5^, which show the limited impact of the Covid-19 epidemic on mortality in France.

## Materials and Methods

### Mortality figures in France

#### Populations and medical causes of death

In France, the statistical death registration system is managed by the National Institute of Statistics and Economic Studies (L’Institut national de la statistique et des études économiques - INSEE)^6^. Death notices are sent out by the municipalities within one week. However, these notices do not specify the causes of death, which are indicated separately in a “confidential medical certificate of death” that must be filled out and signed by a physician and subsequently sent out to the CépiDc anonymously^7^.

Medical causes of death are labeled according to the International Classification of Diseases (ICD-10 version 19) of the World Health Organization (WHO). It is designed to allow comparisons for large groups of causes (e.g., tumors, external causes). An ICD-10 label is assigned to each medical condition (e.g., disease, trauma) mentioned on the certificate. The initial cause of death is then defined as the disease, or the circumstances, that led to the disease process that resulted in death.^8,9^ In view of the outbreak of the Covid-19 epidemic, as of January 2020, WHO produced specific recommendations on how to label the causes of death mentioning the SARS-CoV-2 infection, as well as rules for identifying Covid-19 as the initial cause of death^10,11,12,13^.

To date (February 2023), cause-specific death data from 1979 to 2017^14^ published by the CepiDc are available online to the general public. Data for the year 2020 are not available, but aggregated data are available on the website of the *Direction de la recherche, des études, de l’évaluation et des statistiques* (DRESS) ^15^. These data are sufficient in the context of this study.

#### Estimating excess mortality

To assess the impact of a health event on mortality, it is necessary to estimate the excess mortality during the event, i.e., to calculate the difference between the number of deaths observed the year of the event and the deaths expected for that year should the event not have occurred. The standard method for estimating the expected number of deaths is to consider that it is equivalent to the average number of deaths observed in previous years, in this case the last five years, for equivalent populations, i.e., standardized to the year of the study.

#### Standardization of mortality data

Standardization consists in estimating the number of equivalent deaths in a year according to the age structure so that it corresponds to the same distribution as the one studied, in this case, the 2020 distribution. It allows for a comparison of mortality regardless of changes in the population structure. As of 1 January, INSEE has published population age pyramids by sex and age from 1991 to 2022. We used these data to determine the average populations for each year that produce the number of deaths counted at the end of the year and standardize deaths to 2020.

#### Cause-specific mortality data are missing for 2018 and 2019

2017 is the latest fully labelled year by CepiDc available to date. Consequently, the data for 2018 and 2019 were not readily available to study the year 2020. We are using the method described below to overcome the difficulty associated with this lack of data so as to assess excess mortality without time discontinuity. As a reminder, the problem is to study the impact of the Covid-19 epidemic on mortality in 2020, but the epidemic could not have had an impact on mortality in the years prior to the event. We consider that these years retained the trends that were observed over more than 10 years. Linear regressions were used for the five previous years (from 2013 to 2017). They partially remove the bias caused by these missing data to estimate the expected number of deaths by cause for the year 2020.

#### Scope of the study

The scope of the study includes people residing and deceased in metropolitan France and its overseas departments and territories. In this article, the results are presented for the most frequent categories of causes with numbers greater than 10,000 deaths, to which a specific “Covid-19” category has been added.

For our analyses, we divided the population into three age groups: 1) the group of individuals who died aged under “ 65 years,” 2) those whose age at death was “between 65 and 84 years of age,” and 3) those dead aged “85 years or older”. This choice is imposed by the data made available by the DRESS for the year 2020. It is justified by the sensitivity of each of these groups to health events. The first group corresponds to a young and/or economically active population and is not very sensitive to epidemics in terms of mortality. The second group is that of the elderly, at which age health is a concern. The last group is the elderly, who are particularly sensitive to health events.

## Results

### Mortality by principal causes, all ages and all sexes

In 2020, 667,496 deaths of individuals domiciled in metropolitan France and its overseas departments and territories were recorded in the CépiDc database for an average population of 67,740,259 for that year.

Figure 1 (above) shows the evolution of the first five causes (standardized) for the years 2004 to 2020. The two leading causes of death in France are tumors and cardiovascular diseases. They alone account for about half of all deaths. This is the case since 1979 which is the oldest year available online for deaths by cause.

**Fig. 1:**
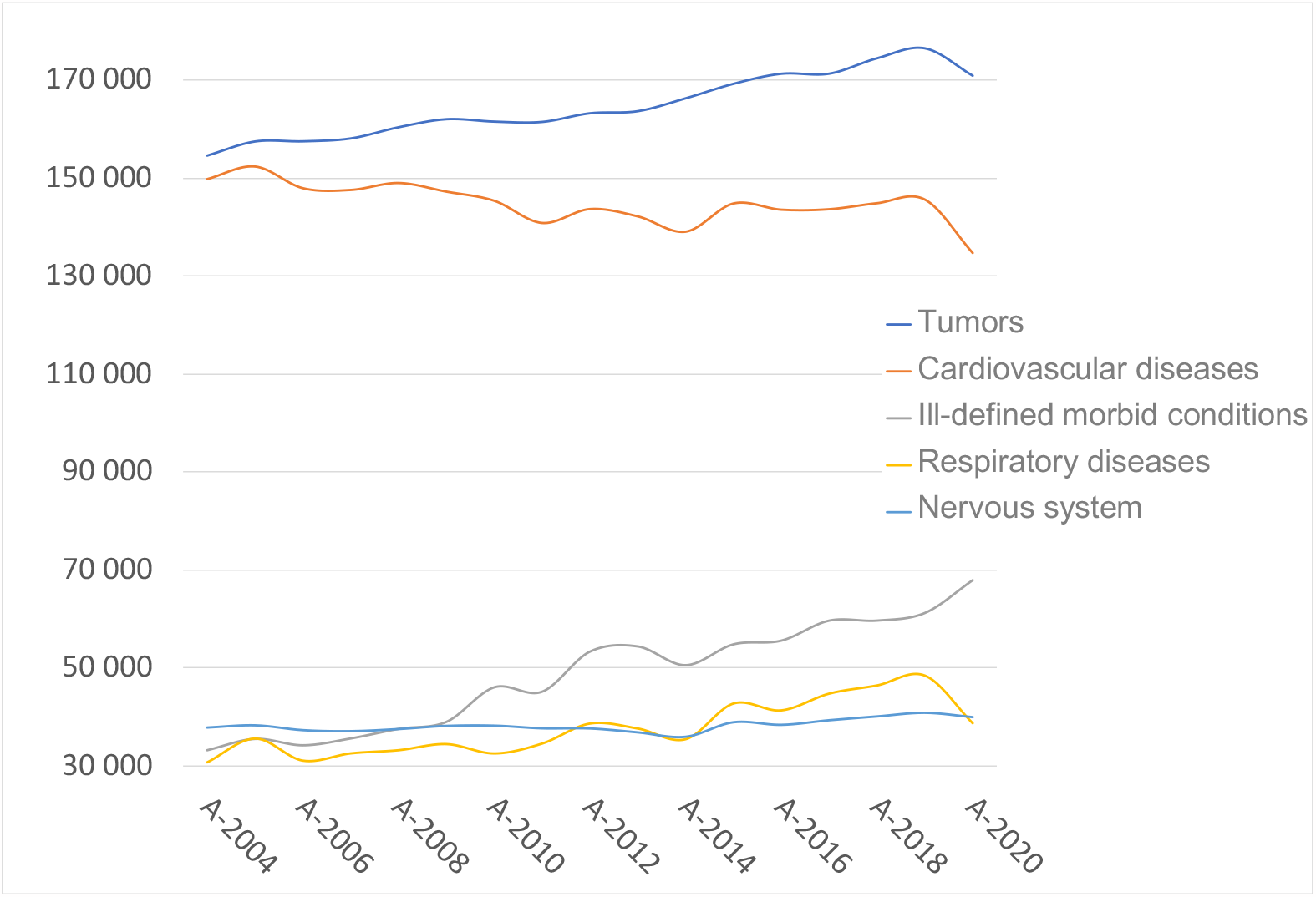
Annual change in the five leading causes of death in France from 2004 to 2020. Source: CépiDc, calculations and formatting by the authors Field: Metropolitan France and its overseas departments and territories Note for the reader: Change in the annual trend in the five leading causes of death since 2004, the year in which tumors became the leading cause of death, overtaking cardiovascular diseases. These two causes alone account for more than half of all deaths for all causes. Tumors are increasing in number but their share among the causes of death is decreasing slightly. In contrast, cardiovascular diseases show a decreasing trend. The next three causes show numbers that are three times lower than the first two.

“Tumors” overtook “cardiovascular diseases” as a primary cause of death for the first time in 2004 and this trend has only been increasing since. Paradoxically, we will see later in this study that, although the statement is correct for the whole population, it is not for 49% of deaths, i.e., the individuals who died older than 85 years of age, for whom “cardiovascular diseases” are still the primary cause of death. This finding is important for the subsequent paragraphs since the concept of causes hierarchy must necessarily consider the age structure to avoid misinterpretations.

### The introduction of “Covid-19” as a cause of death changed the 2020 ranking

Since 2004, the ranking and trends of the leading causes of death are stable. All causes are constantly increasing. This is due to the global increase of the population in France and in particular with regard to the number of elderly people. 69,238 deaths were labeled as “covid-19”, i.e., 10.4% of the deaths recorded in 2020.This number may appear significant, but it remains modest compared with that of “cardiovascular diseases”, which comes ahead of it in the ranking with 134,763 deaths (i.e., almost twice as many). Nevertheless, this figure places this new cause in third position, just before the 67,772 deaths labeled as “ill-defined morbid conditions”.

Figure 1 shows an abnormal break in the trend for the year 2020, particularly for the first two causes. The following paragraphs describe this evolution.

#### Evolution of the first two causes which account for more than half of the total deaths

As described above, the cause of death “tumors” has been the leading cause of death since 2004 with a share of 29.9% of deaths that year. Since then, while the number of deaths from this cause has been increasing (especially since 2013) (see Fig. 2), its share among all causes has been decreasing slightly but steadily by 0.11% per year on average. Strangely in 2020, the year of the first Covid-19 outbreak, the percentage of tumors dropped to 25.6% or -2.7pts compared to 28.3% in 2017, which is the latest year with available data. Consequently, 174,062 deaths from “tumors” were expected but only 170,806 were observed, which represents a statistically significant shortfall of about 3,256 deaths. Such a drop is abnormal for chronic conditions that are not directly impacted by a communicable disease epidemic such as Covid-19.

**Fig. 2:**
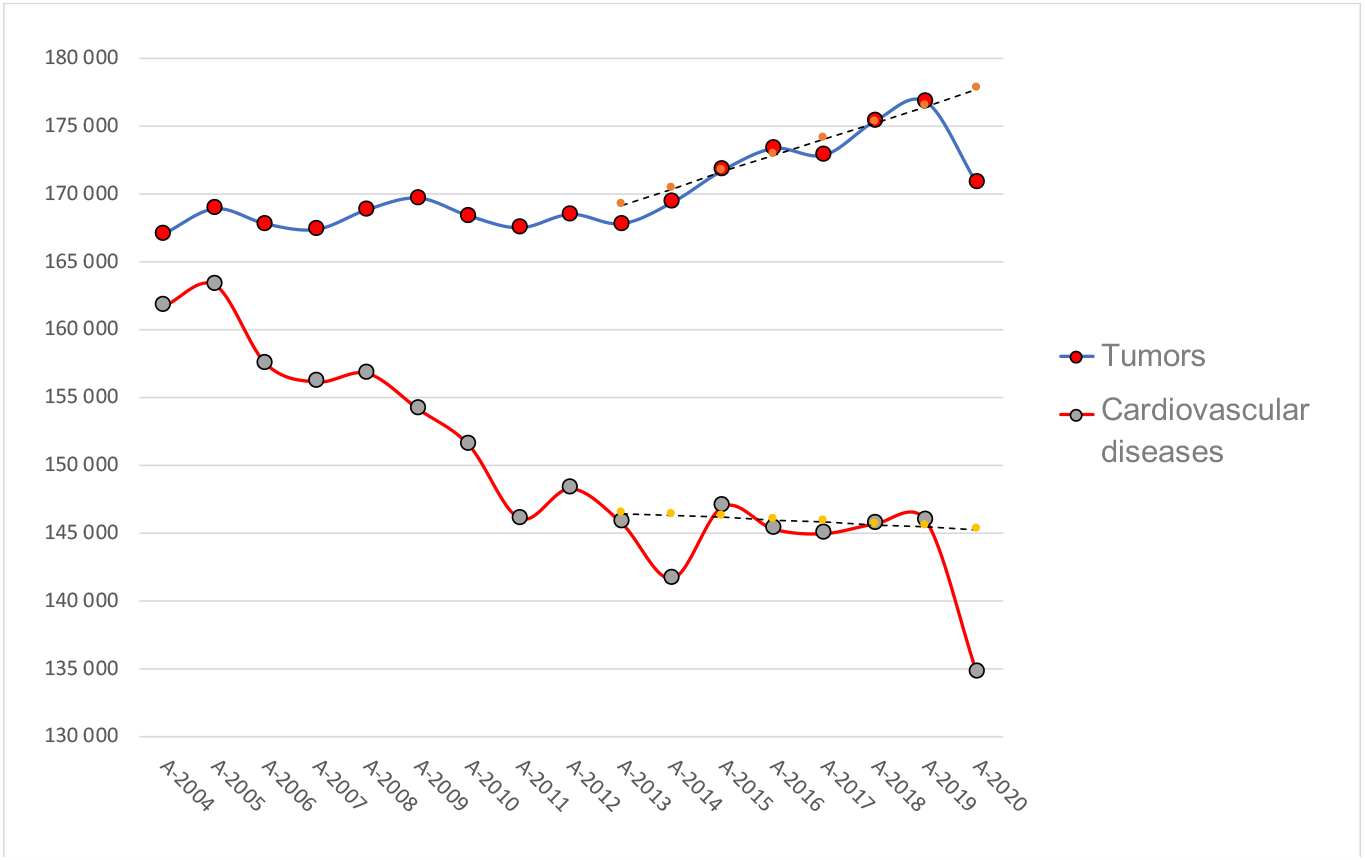
Standardized annual change in the two leading causes of death in France from 2004 to 2020. Source: CépiDc, calculations and formatting by the authors Field: Metropolitan France and its overseas departments and territories Reading note: Change in the annual trend of the first two causes of death standardized to 2020. The two years 2017 and 2018 are estimated by linear regression on the previous five years. The trend line for these causes is dotted with an estimated value for 2020 to be compared to the observed declining value in 2020

Cardiovascular diseases were the second most common cause of death. As previously stated, until 2004, it was the leading cause of death in France. In that year, they still accounted for 28.9% of deaths. Since then, this percentage has steadily decreased by 0.37pts annually. In 2020, the year of the first covid-19 outbreak, the percentage of cardiovascular diseases fell sharply to 20.2%, i.e., -3.6pts compared with 23.8% in the latest reported year three years earlier (see Figure 2). 145,828 deaths from this cause were expected but only 134,763 were observed, which represents a statistically significant shortfall of 11,065 deaths. Such a drop, as was observed for tumors, raises questions. In no way can such an observation be considered a protective effect of the epidemic, which would be tantamount to inferring that an epidemic of communicable disease would lead to a reduction in tumors and cardiovascular diseases, a far-fetched possibility. We will show that the causality is more prosaic.

#### Trends in the next three causes which account for a quarter of all deaths

The next three causes have numbers of deaths 3 to 4 times lower than the first two (see Fig. 1). The cause “ill-defined morbid conditions” comes in third place with 9.6% of deaths on average since 2015. This is the only cause with many deaths that shows an increase against the expected. 58,594 deaths from this cause were expected and 67,772 were observed, which represents an excess of 9,178 deaths.

“Respiratory diseases” come fourth with 7.4% of deaths on average since 2015. For this cause, 45,146 deaths were expected but only 38,711 were observed in 2020, a significant shortfall of 6,435 deaths. While a disease affecting the respiratory tract was rampant, the cause “respiratory diseases” has seen its numbers drop. Such a drop would be misinterpreted had we not considered the bias introduced by the specific implementation of “Covid-19” as a cause of death.

Diseases of the nervous system come in fifth place with 6.6% of deaths. The share of this cause has been increasing strongly and constantly for more than 30 years, but in 2020 it also decreased compared with the previous year. Here again, such a drop is difficult to explain because this type of pathology is not directly linked to an epidemic of a transmissible disease. It is probably a transfer from this cause to the “Covid-19” as a cause of death.

#### Changes in the following causes

The causes that follow have been responsible for less than 7% of deaths on average since 2015. In descending order, we find the external causes with 6.5% of deaths, mental disorders with 4.4% of deaths. Regarding the trend, in 2020, we again observe a significant decline, as we did for diseases of the nervous system (see Table 1). Next come diseases of the digestive system with 4.0% of deaths, followed by endocrine diseases with 3.7% of deaths and infectious diseases with 1.8% of deaths.

**Table 1:** Assessment of the gap between expected and observed mortality in 2020 by cause of death.

| Cause of death | Expected number of deaths in 2020 | Observed number of deaths in 2020 | Deviation from expected | Rate against the expected | Significant |
| --- | --- | --- | --- | --- | --- |
| Tumors | 174,062 [172,275 ; 175,850] | 170,806 | -3,256 [-5,044 ; -1,469] | -0.53% | Yes |
| Cardiovascular diseases | 145,828 [145,161 ; 146,494] | 134,763 | -11,065 [-11,731 ; -10,398] | -1.80% | Yes |
| Ill-defined morbid conditions | 58,594 [56,344 ; 60,844] | 67,772 | 9,178 [6,928 ; 11,428] | 1.49% | Yes |
| Respiratory diseases | 45,146 [42,826 ; 47,467] | 38,711 | -6,435 [-8,756 ; -4,115] | -1.04% | Yes |
| Nervous system | 39,842 [39,020 ; 40,664] | 40,033 | 191 [-631 ; 1,013] | 0.03% | No |
| External causes | 40,379 [39,231 ; 41,527] | 37,616 | -2,763 [-3,911 ; -1,615] | -0.45% | Yes |
| Mental disorders | 26,939 [25,884 ; 27,994] | 25,388 | -1,551 [-2,606 ; -496] | -0.25% | Yes |
| Digestive system | 24,630 [24,361 ; 24,900] | 24,975 | 345 [75 ; 614] | 0.06% | Yes |
| Endocrine diseases | 22,346 [21,779 ; 22,912] | 23,598 | 1,252 [686 ; 1,819] | 0.20% | Yes |
| Infectious diseases | 11,302 [10,952 ; 11,651] | 11,055 | -247 [-596 ; 103] | -0.04% | No |
| genitourinary system | 15,506 [10,508 ; 20,504] | 12,092 | -3,414 [-8,412 ; 1,584] | -0.55% | No |
| Sum of other causes | 11,619 [11,343 ; 11,895] | 11,449 | -170 [-446 ; 106] | -0.03% | No |
| Covid |  | 69,238 | 69,238 | 11.24% |  |
| **Total** | **616,194 [609,652 ; 622,735]** | **667,496** | **51,302 [44,761 ; 57,844]** | **8.33%** | **Yes** |

Source: CépiDc, Insee Demographic Indicators, authors' calculations.
Field: Metropolitan France and its overseas departments and territories
Note for the reader: This table groups the results of the mortality analysis in 2020 by cause of death (all ages and all sexes) sorted by decreasing order for each cause, except for "Covid-19" which is placed last because it was only observed in 2020. The column "Expected number of deaths in 2020" is the average of the deaths observed in the previous five years for populations equivalent to that of 2020. The results of this estimate are presented with their 95% confidence intervals. The "Deviation from expected" column is the difference between expected and observed deaths. The rate of increase over the expected is in the next column. The last column indicates the significance of the result.

These first ten causes account for 96.3% of all deaths in France on average over a 20-year period. The next seven causes account for less than 10,000 deaths per year. For clarity purposes, we have grouped them together under the “sum of other causes” heading in this analysis.

The following table groups the results of this paragraph on the analysis of mortality in 2020 by causes (all ages and all sexes) sorted in descending order for each cause (except for “Covid-19,” which was only observed in 2020 and was placed last to be easily singled out).

We note that, out of the twelve major causes of death, eight have seen their numbers decrease, five of which significantly. The cumulative excess of causes of death shows 17,936 fewer deaths than expected. The shortfall in deaths for all these causes seems to have been transferred to Covid-19 as a cause of death, making it third in the ranking. Consequently, this new cause of death was heavily used as a label while, at the same time, most other causes witnessed a significant drop in a context in which the total number of deaths did not increase in the proportions announced. In the following paragraph, the analysis of deaths by age group sheds additional light on this issue.

### Age-specific mortality analysis using CepiDc data

In a previous study using INSEE data, the population was divided into two groups at age 65. The distribution of these two categories verified Pareto’s law^16^ with more than 80% of deaths coming from the 20% of the population over 65 years of age. This finding is strengthened if we consider that almost half of the deaths (49%) came from the 3% of the population aged over 85 years. The causes of death in this age group are not the same as those of the rest of the population. This category of the population is more fragile and therefore more vulnerable to health events such as a heat wave or an epidemic. Consequently, a health event will have important consequences on the analysis of the hierarchy of causes because the number of deaths is very high in this advanced age group, which is particularly fragile, compared to the rest of the population. This age group also differs in that certain diseases that mainly affect other age groups, such as those labeled as “tumors”, no longer play a significant role. These considerations require an analysis by age group.

Figure 4 (above) shows the disproportions in population size and deaths for the three age categories chosen for this study (this choice is explained above in the Materials and Methods section). Such disproportions call for caution when studying the real impact of a risk factor, such as a communicable disease epidemic, on a population.

The CépiDc database indicates 667,496 all-cause deaths in 2020. The expected number of deaths excluding Covid-19, is estimated at 616,194 [609,652; 622,735] deaths. Consequently, during 2020 with the Covid-19 epidemic, there were allegedly 51,302 [44,761; 57,844] additional deaths. These additional deaths give an overall excess mortality of 8.3% [7.3%; 9.4%]. However, the Yule-Simpson statistical effect^17^ due to differences in proportions between age categories must be considered in order to find an estimate that accounts for the differential in numbers for the three age groups. This results in an excess of deaths of 27,775 [15,718; 39,838] individuals in 2020, i.e. an effective excess mortality of 4.3% [2.5%; 5.9%] (see Table 2). These results obtained with data from the CépiDc are consistent with those obtained in our study based on the INSEE as early as March 2020^18^.

**Table 2:**
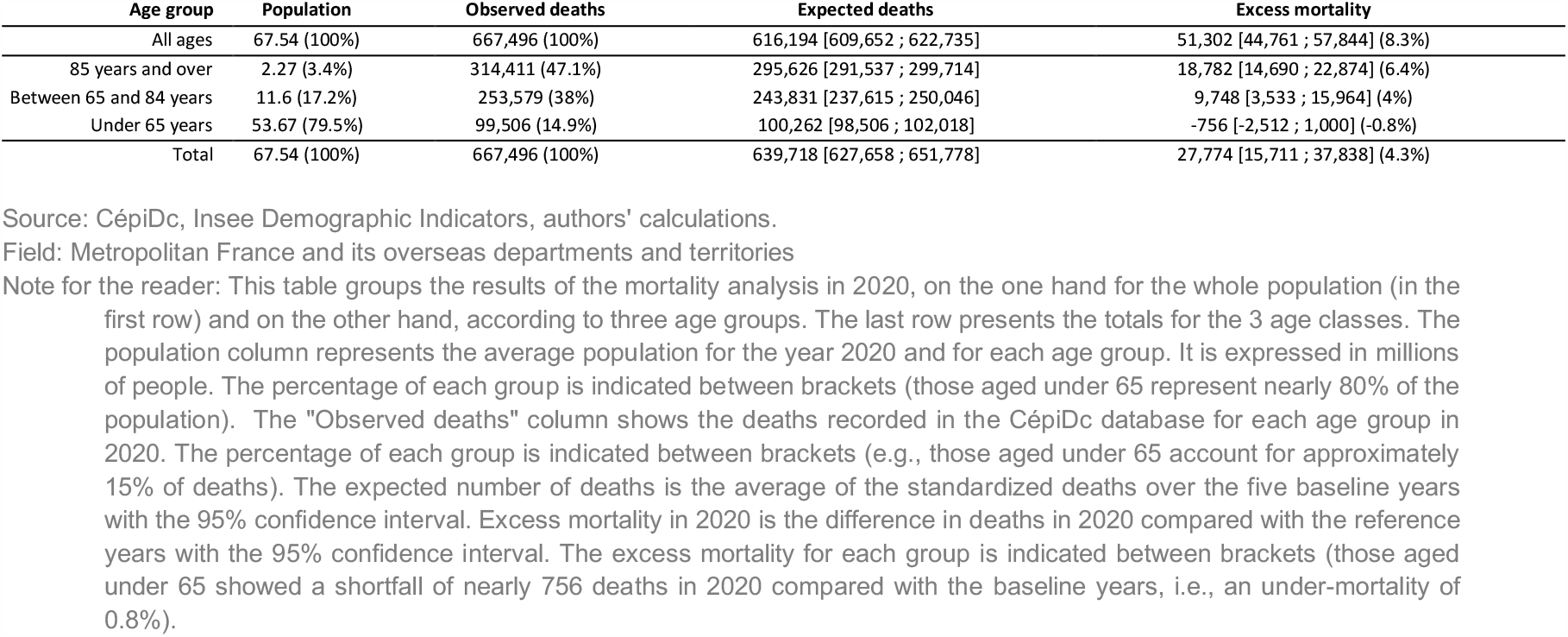
Assessment of the gap between expected and observed mortality in 2020 by three age categories.

The figure below makes it possible to compare the impact of the different causes regardless of the numbers in the groups studied, because the number of deaths for each of them is standardized to a population of 10,000 in the group. It appears very clearly that the group aged over 85 outnumbers the others, in particular for the cause “cardiovascular diseases” (in 2^nd^ place) but this is the case for all causes and this calls for a detailed study for each group.

### Results for deaths that occurred at age “under 65 years”

In 2020, only 99,506 deaths out of the 667,496 recorded in the CépiDc database (i.e.,14.9%) were under 65 years of age, whereas this age category represents 79.6% of the population (53.7 million individuals). This is an obvious observation since it is generally accepted that there is a low probability of dying young in France in our time. The other observation is that this percentage has been constantly decreasing for at least three decades, which means that deaths are occurring at an increasingly advanced age and 2020 was no exception.

For this age group, the difference between observed and expected deaths was -756 [-2,512; 1,000], i.e., a negative excess mortality (under-mortality), which was not significant but estimated at -0.7%. Consequently, despite the epidemic, there was no excess mortality for nearly 80% of the population. However, the same calculations of excess mortality over a 20-year period show a chronic under-mortality for this age group except for 2 years, 2009 and 2010, with a 0.6% and 0.2% excess mortality respectively. As such, over a 20-year period, the only years in which this age group shows excess mortality correspond to the H1N1 epidemic but not to the Covid-19 epidemic. This finding seriously challenges the catastrophic perception of the Covid-19 epidemic.

“Tumors” is by far the leading cause of death for people under 65, as it is in the overall population. Since 2004, this cause has been responsible for over 40% of deaths for this age group. In 2020, the share of this cause among all causes fell by 3.3%. We must go back to 1995 (25 years) to find a figure lower than that of 2020.

The introduction of “Covid-19” as a new cause of death in 2020 explains this drop. The number of 4,301 Covid-19 labeled deaths for this age group is roughly equivalent to the shortfall of 3,475 deaths for the cause “tumors” against the expected in 2020. In addition, among the twelve leading causes, nine showed an under-mortality against the expected, eight of them significantly (see Table 3). The result shows a balance between what was attributed to the Covid cause and the shortfall observed in all other causes.

**Table 3:**
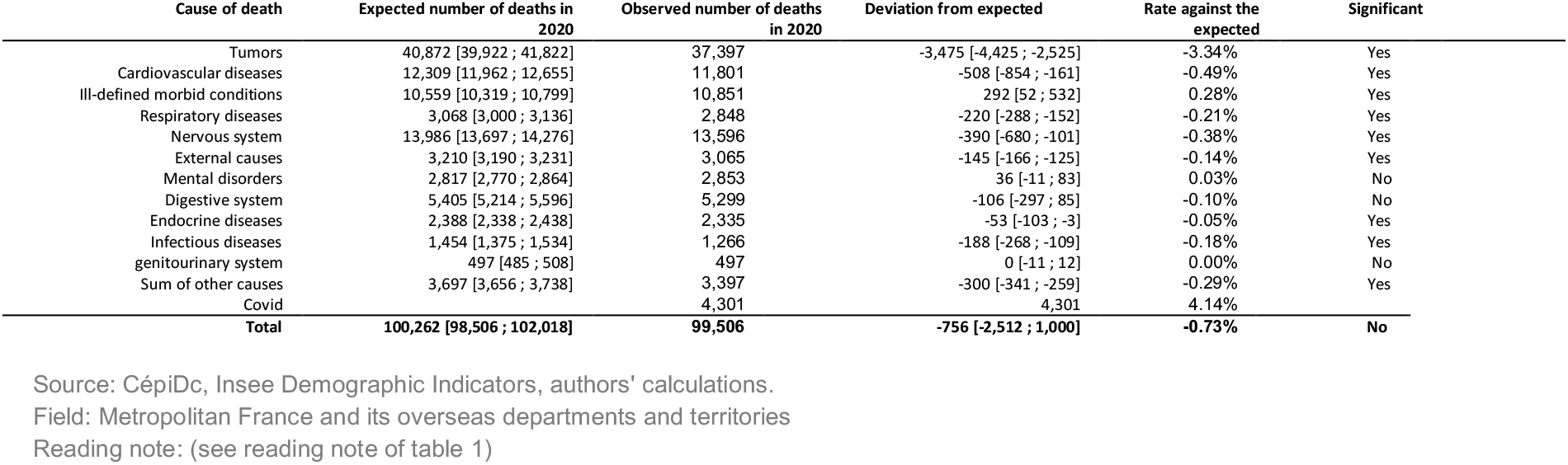
Assessment of the gap between expected and observed mortality in 2020 for “under 65”.

**Table 4:**
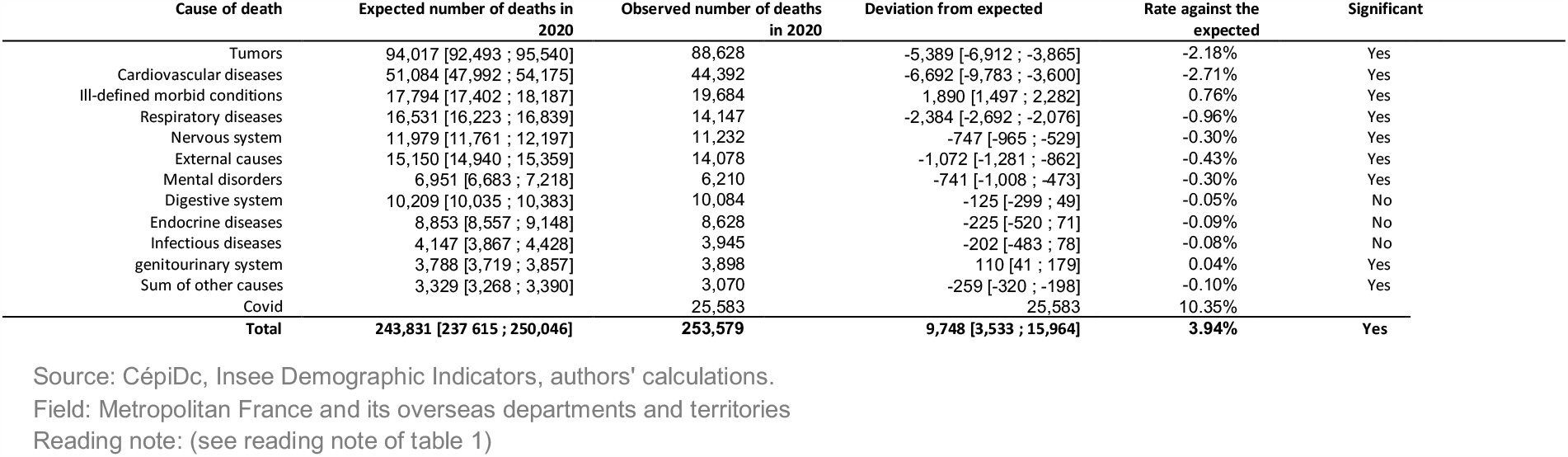
Assessment of the gap between expected and observed mortality in 2020 for population aged “between 65 and 84 years”.

**Table 5:**
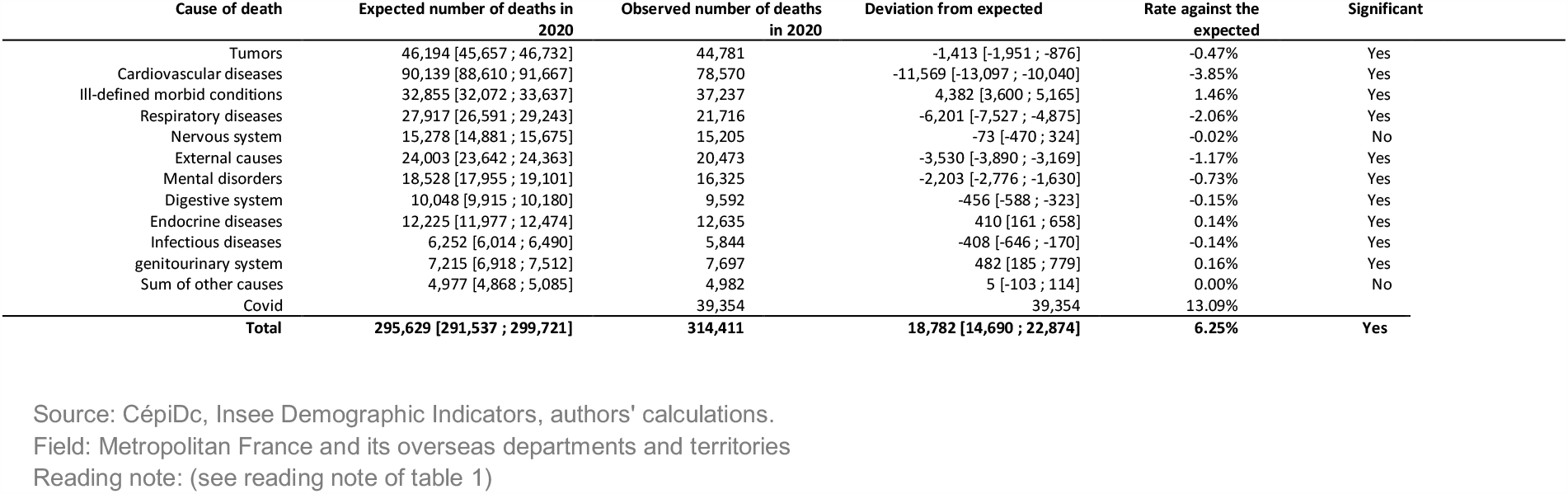
Assessment of the gap between expected and observed mortality in 2020 for the “85+” age group.

For this age group, the ranking is different from the one presented in the previous paragraph for the general population. The second cause is not “cardiovascular diseases” but external causes (among which accidents are listed); this cause has not witnessed any significant changes, as it shows a shortfall of 390 deaths against the expected, i.e., an under-mortality of 0.06%. This result is interesting because it shows that, unlike what was announced, the lockdowns and the associated limitation of movements did not have a protective effect on the number of accidents. In fact, the exceptional introduction of a new cause, “Covid-19”, places it in 6th place with 4.3% of deaths for this age group, whereas the great majority of the other causes have seen their numbers drop against the expected (cf. Table 3).

### Results for deaths that occurred at ages “between 65 and 84 years”

This age group represents 17.2% of the population. In 2020, 253,579 (38%) deaths occurred between the ages of 65 and 84. This large number of deaths for an age group with a relatively small population indicates that this age group corresponds overall to the average life expectancy of the French population. It is thus expected that many deaths will occur within this age group. Compared to the expected deaths excluding Covid-19, 9,748 [3,533; 15,964] additional deaths were counted, i.e., an excess mortality of 3.94%. Without being exceptional, this excess mortality is the sign of a significant health event, even though not uncommon. It is undeniable that the increase in life expectancy has played a role in the fact that this age group has shown a systematic under-mortality for the last twenty years. We must go back to 2003, the year of the heat wave, to observe an almost equivalent excess mortality of 3.2%. consequently, the “2003 heat wave” health event, which lasted only a few days, had an effect equivalent to that of the Covid-19 epidemic, which lasted many months. This raises questions about the overestimation of its impact during this period.

For this age group, the ranking of causes of death is the same as for the general population, except for mental disorders, which are less frequent for this age group unlike for individuals over 85 years of age.

As for the under 65s, 2020 is exceptional in that ten out of twelve causes showed an under-mortality (seven of them significantly) against the expected without “Covid-19”. Although the cause “covid-19” has a headcount of 25,583 deaths, the sum of death shortfalls for the other causes amounts to 17,835 deaths short of what was expected. One of the few causes with an excess mortality against the expected is “ill-defined morbid conditions” with an excess of 1,890 additional deaths. In the discussion, we will explain this excess, which may appear abnormal.

### Results for deaths at age “85 years and over”

314,411 (47.1%) of the deaths were older than 85. This age group represents only 3.4% of the population. However, its size has more than doubled in 30 years, with an increase rate of 3% per year. Compared to expected deaths outside the epidemic, 18,782 [14,690; 22,874] additional deaths were recorded, i.e., an excess mortality of 6.25%. Unlike for the previous age groups, excess mortality is recurring for this age group. This can be explained by the fact that life expectancy is not infinite, and individuals eventually die. This usually happens within this age group. 6.4% can be regarded as a high excess mortality, even though not exceptional. In 2003, the year of the heat wave, the excess mortality for this age group was 10.0%. Here again, the “2003 heat wave” health event, which lasted only a few days, had a much greater impact than the Covid-19 epidemic.

The particularity of this age group is that the cause “tumors,” which is by far the first cause of death for all other ages, only comes in second place with about 15% of deaths, far behind “cardiovascular diseases,” which come in first place with about a third of deaths. It turns out that it is also this cause that shows the greatest drop against the expected with an under-mortality of almost 4% with 11,565 fewer deaths compared to those expected in the absence of an epidemic and therefore largely contributes to offsetting the number of deaths attributed to the cause “covid-19”. As for the other age groups, this one also shows an exceptional fall in most of the other causes. The sum of all death shortfalls amounts to 25,852 fewer deaths than expected, while simultaneously, the cause “covid-19” shows 39,354 deaths. As was described for the other two age groups, we observe a form of transfer to this new label set up on an ad hoc basis. Covid-19 is a respiratory disease. However, the “respiratory diseases” cause, which should have been used to label deaths due to this disease, also shows a decrease of more than 2% in 2020.

## Discussion

### Limitations of using causes of death

#### Limitations related to missing data

The CépiDc of Inserm works on the basis of “causes of death” filled in by physicians on death certificates. Raw data are available from 1979 to 2017. For 2020, we used the aggregated data as they were made available on the DRESS website^4^. This exercise comes with many limitations. Since death certificates for 2018 and 2019 were not yet labeled, we estimated the values of the causes for these years using the precautions explained in the “Materials and Methods” paragraph. As the choice of age groups was imposed, it was impossible to respond in greater detail to certain hypotheses pertaining to the consequence of the evolution of the age structure in France for certain risk factors. Similarly, the population residing and deceased in the overseas departments and territories were included in the data, which poses real methodological problems. It would have been interesting to gain access to this information to distinguish between metropolitan France and the overseas territories for obvious reasons of bias due to geographical disparities. Local population characteristics have an influence on mortality patterns, particularly in the context of a communicable disease epidemic while population transfers have been notoriously disrupted. On the other hand, these data have already been analyzed by other authors and have been the subject of publications, notably by the bodies that produced them^19^. It is on this basis that we have used them as they stand, without discussing them, considering them, *a priori*, as reflecting a reality.

#### Perverse effect of diagnostic innovations

However, a quick evaluation of these data shows many inconsistencies. A posteriori reflection is necessary, first and foremost, by questioning the quality of the data. Data from death certificates have the qualities and defects of administrative sources. They cover a very large part of the population. But, unlike surveys, they do not guarantee perfect comparability over time. For example, technological innovations make cancer screening tools increasingly sensitive. It is therefore difficult to distinguish, within the increase in the cancer-labeled cause on death certificates, between what relates to the increase in the disease and what is to be attributed to its detection. The same applies to the introduction of PCR tests for screening and the “covid-19 confirmed” label.

#### Perverse effect of the introduction of the “Covid-19” label

In 2020, in the context of the collection of these data, several important innovations were implemented. The first, and not the least, is the production by the WHO, from January 2020, of a specific label and recommendations for labelling the initial cause of deaths mentioning a SARS-CoV-2^10^ infection. Such an introduction can only be justified by a catastrophic event in terms of excess mortality. However, as early as March 2020, epidemiological analyses already showed that the health impact in 2020 would be limited and equivalent to the usual winter epidemics^20,21,22^. This study on the causes of death provides new elements that confirm these projections. Changes in ICD-10 coding necessarily have an impact on the final results. The introduction of a new specific label in a context of low excess mortality inevitably produces obvious effects on the hierarchy of causes, where transfers between causes occur rather than real increases. This is what we have highlighted in this study.

#### Effect of algorithm-generated labelling on the free texts of certificates

A second innovation is the effective application in 2020 of an algorithm to identify the “Covid” mention in the free text of the medical sections of death certificates. In fact, 77,535 deaths were detected with this mention and almost 90% of them have been labeled with “covid-19” as the initial cause of death. In other words, the mere fact that a “Covid” mention appears in a free text is enough to validate it as the initial cause of death with a probability of 90%. It should be remembered that death certificates generally mention several causes, whether they are placed on the same level or in a hierarchy^23^. In two thirds of cases, death certificates mentioning “Covid-19” as the initial cause of death reveal the presence of known major comorbidities (e.g., heart diseases, arterial hypertension, severe diabetes, renal and respiratory pathologies). The presence of these comorbidities induces a confounding factor that does not allow for death differentiation. It should also be remembered that the “medical death certificate” can be completed by a physician or a postgraduate medical student, regardless of their prior knowledge of the deceased’s health.

#### Compliance in a crisis context and other confounding factors

Consequently, in a context of psychosis, to which most medical personnel were not insensitive, even consciously feeding it with contradictory injunctions or resorting to arguments of authority without scientific basis; in a context in which the prominent discourse was to impute to Sars-Cov2 all sorts of health disorders, even the most far-fetched ones, it is not surprising to see “Covid” mentions on death certificates, and hence in the statistics. A conventional ripple effect, known from experience in the behavior of medical observers in electronic surveillance systems for communicable diseases, consists in attributing the cause of the disease, and in this case of the death, to the epidemic of the moment.

#### What accounts for the increase in “ill-defined morbid condition” and “accidental fall”?

It is extremely rare for an autopsy to be performed on elderly people found dead at home. If the death certificate is not completed by the physician who was treating the person prior to death, the initial cause of death is unreliable in a large proportion of cases and is filled in arbitrarily by the physician certifying the death. During such a troubled period, in a context of incitement, the “Covid” label fulfilled this role, as was to be expected and was confirmed by this study. This is probably also the reason why one of the few causes that witnessed an increase in 2020 was the cause “ill-defined morbid condition”. This increase indicates that a significant number of deaths that usually occurred in hospitals occurred at home, thereby suggesting a lack of care.

For the record, throughout 2020, hospital activity dropped by 10% compared to previous years due to deprogramming^24^. Covid-19-related activity only accounted for 2% of total activity for the year^25^. On the other hand, the increase in deaths due to accidental falls among people aged over 85 years could reflect the lack of care for this age group.

#### How an administrative decision on a drug shortage changed labeling

A third issue concerns an administrative rule implemented in March 2020 that may have had a significant impact on the reporting of covid-19-labeled death data. The beginning of 2020 was marked by a worldwide shortage of Midazolam^26^, a drug used in palliative care. The French government then authorized Rivotril^27^ by decree as a replacement drug, but only for “patients affected or likely to be affected by the SARS-CoV-2 virus whose clinical condition justifies it upon presentation of a medical prescription bearing the mention “Off-label prescribing in the context of covid-19 (“*Prescription hors AMM dans le cadre du covid-19*”). The word “likely” highlights the limited information available to physicians before using this prescription. On the other hand, this decree may have had a significant impact on cause-of-death statistics: if the physician considers that the patient’s condition requires the injection of a palliative product, they must declare the prescription as “within the framework of covid-19”, which resulted in a new record of death for this disease. The administrative decision with regard to a drug shortage consequently affected death labeling.

### The causes of death show the minor impact of the Covid-19 epidemic

Studying the evolution of general mortality over several years, analyzing it by age group and reasoning in terms of excess mortality can be judicious to circumvent the labeling bias mentioned above. A few weeks into 2021, several articles already cited answered this important question based on vital statistics and population estimates released by INSEE. The excess mortality in France for the year 2020 has been estimated at between 25,000 and 58,000 additional deaths depending on the methodology used. An INSEE study estimates 47,000 excess deaths from all causes against the expected^28^. Let us keep this figure to avoid any unnecessary controversy in this case.

Logically, if the WHO decides to set up a specific “Covid-19” label, it is because it expects that the number of deaths related to this disease will be greater than the expected and therefore that the excess mortality will be significant and at least equal to the number of deaths identified with the cause of this disease. However, the CépiDc counts 69,238 covid19-labeled deaths. This means that the excess mortality shows 22,000 fewer deaths (-47%) than the number of covid19-labeled deaths in 2020. In other words, if the number of covid-19-labeled deaths exceeds the excess mortality, the additional deaths for this cause must come from elsewhere. This means that the difference would have been attributed to other causes in the absence of this cause set up in a particular context. This is exactly what our study on cause-specific mortality data shows.

#### No excess mortality for 80% of the population and a small excess mortality for the remaining 20%, is this serious?

The other argument that confirms the limited impact of the Covid-19 epidemic is the fact that 80% of the population, the under 65s, have not shown any excess mortality in 2020. It is worth remembering that, for the 53.7 million population in this class, only 4,301 deaths were labeled as “Covid-19”. We have shown (see above: Results for deaths at age “under 65 years”) that the H1N1 epidemic was more deadly than the Covid-19 epidemic for this age group. In the same way, the other studied classes of older individuals show that the “Heat wave 2003” event, which also affected only the elderly, had higher excess mortality without having had the societal consequences brought about by the Covid crisis.

#### An exceptional decrease in most causes of death

Whether for the population as a whole or by age group, mortality in 2020 stood out not by its excess but on the contrary, the abnormal decrease in deaths observed for almost all causes of death apart from those labeled with Covid-19. We have shown that mortality due to tumors and circulatory system diseases, which account for the first two main categories of causes of death, had decreased significantly in 2020, regardless of the age group (see fig. 2). Consequently, the number of missing deaths is balanced out by the number of deaths attributed to Covid-19 for the “under 65” age group.

For other age groups, the pandemic has probably triggered the death of elderly or frail people with comorbidities. However, such a transfer of a set of causes to the new cause “Covid-19” necessarily indicates a labeling bias. Conversely, and as we have already mentioned, the continuing upward trend in mortality due to accidental falls among the elderly may have been fostered by the isolation brought about by the lockdowns and the resulting reduction in contacts decided by the health authorities to limit the contamination of vulnerable people, which led to abandonment and therefore a lack of care.

#### In France, in the midst of a respiratory syndrome epidemic, the cause “respiratory diseases” decreased

In France, “respiratory diseases”, with 7.4% of deaths on average, come 4th in the hierarchy of causes of death. In 2020, this cause of death fell dramatically (see Fig. 3) while a disease affecting the respiratory tract was rampant. For this cause, even more than for others, it is evident that a form of transfer has been operated to another more specific cause such as “Covid-19”, thereby artificially inflating its prevalence. This specification does not denote an extraordinary severity of the disease. The decrease in respiratory diseases is still a French exception, unlike Italy or the United States, which report an increase in deaths from respiratory diseases.

**Fig. 3:**
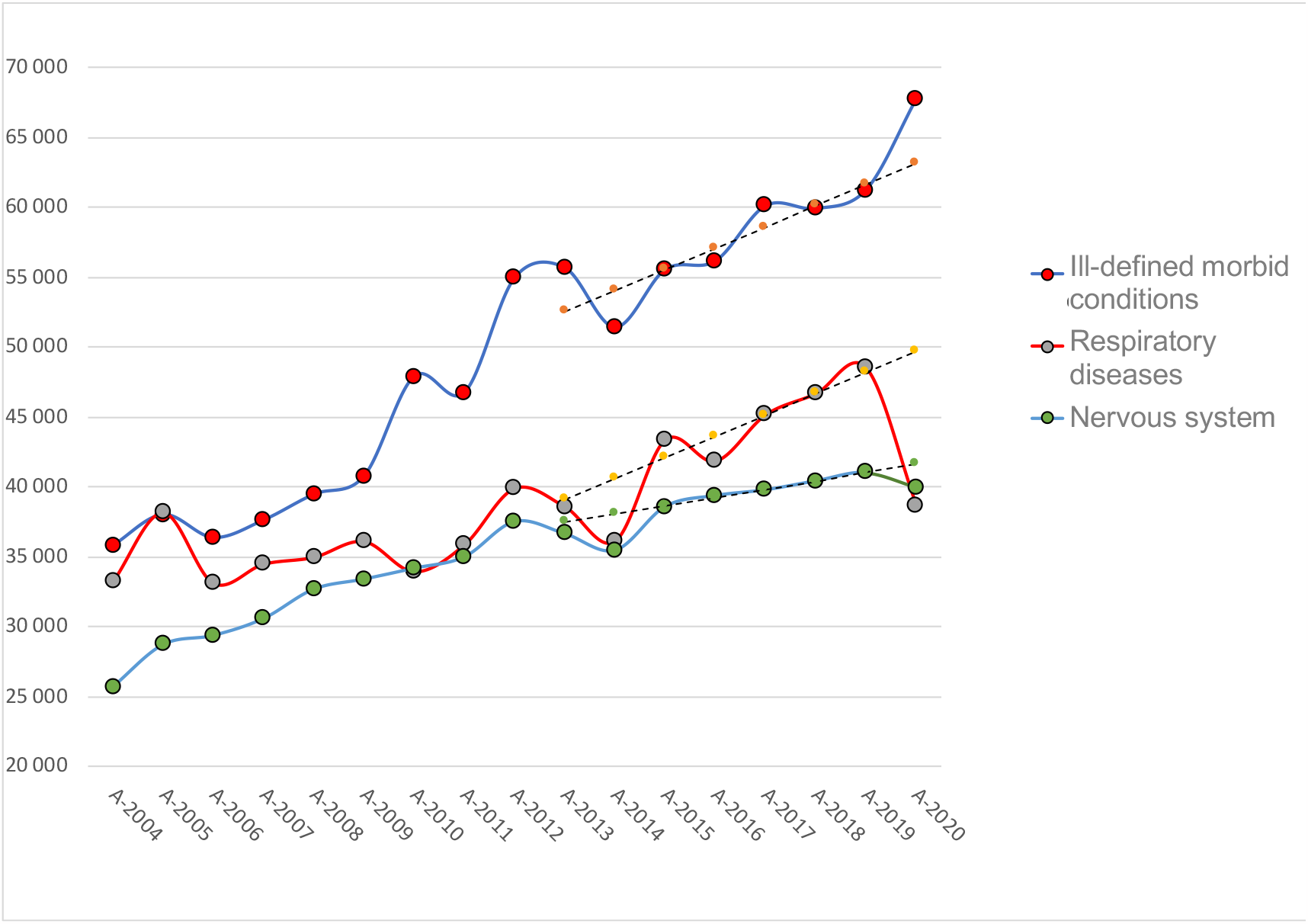
Standardized annual change in the following three causes of death in France from 2004 to 2020. Source: CépiDc, calculations and formatting by the authors Field: Metropolitan France and its overseas departments and territories Reading note: Change in the annual trend of the next three causes of death standardized to 2020. 2017 and 2018 are estimated by linear regression on the previous five years. The trend line for these causes is dotted with an estimated value for 2020 to be compared to the values observed in 2020. Respiratory and nervous system diseases are falling back to the low 2014 values. The cause “Ill-defined morbid conditions” is one of the few causes to go over the trend line.

**Fig. 4:**
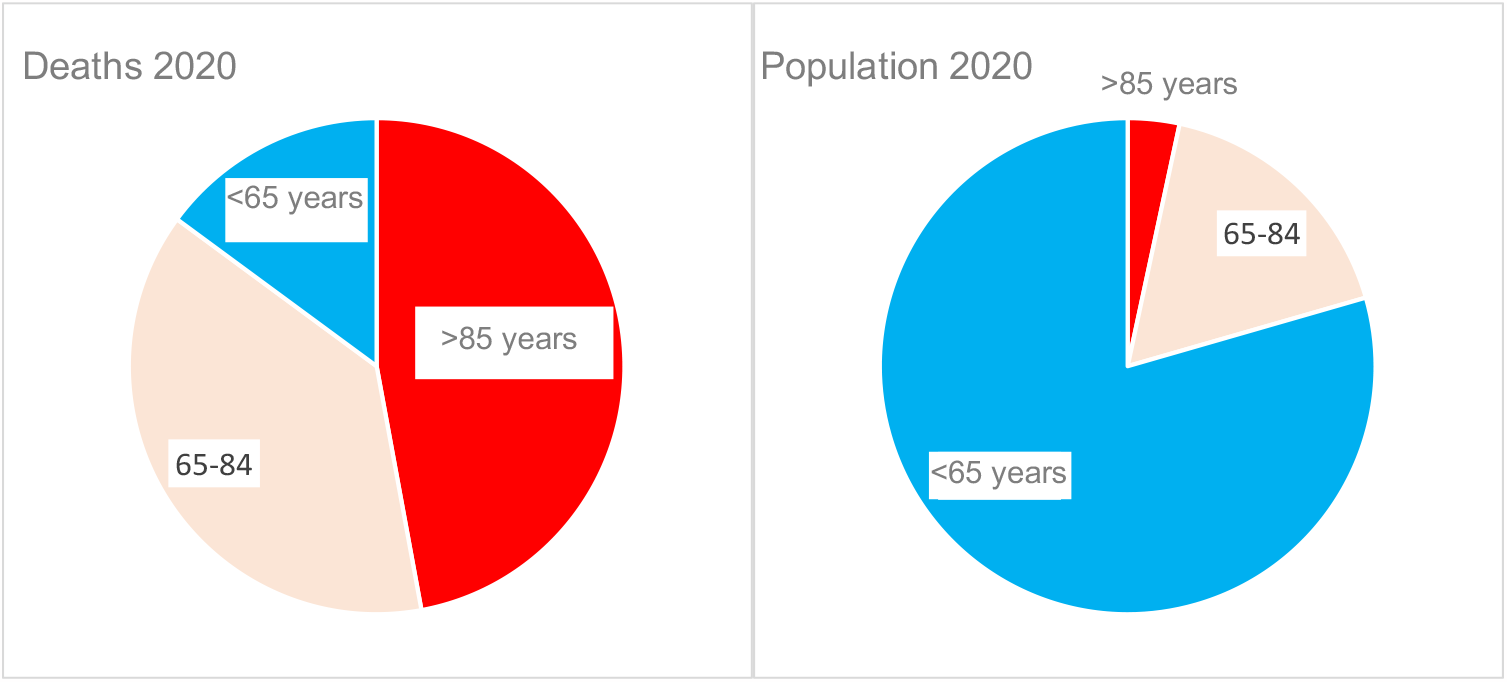
Share of each age group in the study for population and deaths for the year 2020. Source: CépiDc, calculations and formatting by the authors Field: Metropolitan France and its overseas departments and territories Note for the reader: The study focuses on three age groups: 1) those “under 65” (blue), 2) those “between 65 and 84” (pink), and 3) those “over 85” (red). These two graphs highlight the effect of age on mortality in terms of numbers, on the one hand on populations and on the other hand on deaths. The disproportions are graphically obvious, for example in the red areas nearly half of the deaths (49%) come from the 3% of the population aged over 85 years.

**Fig. 5:**
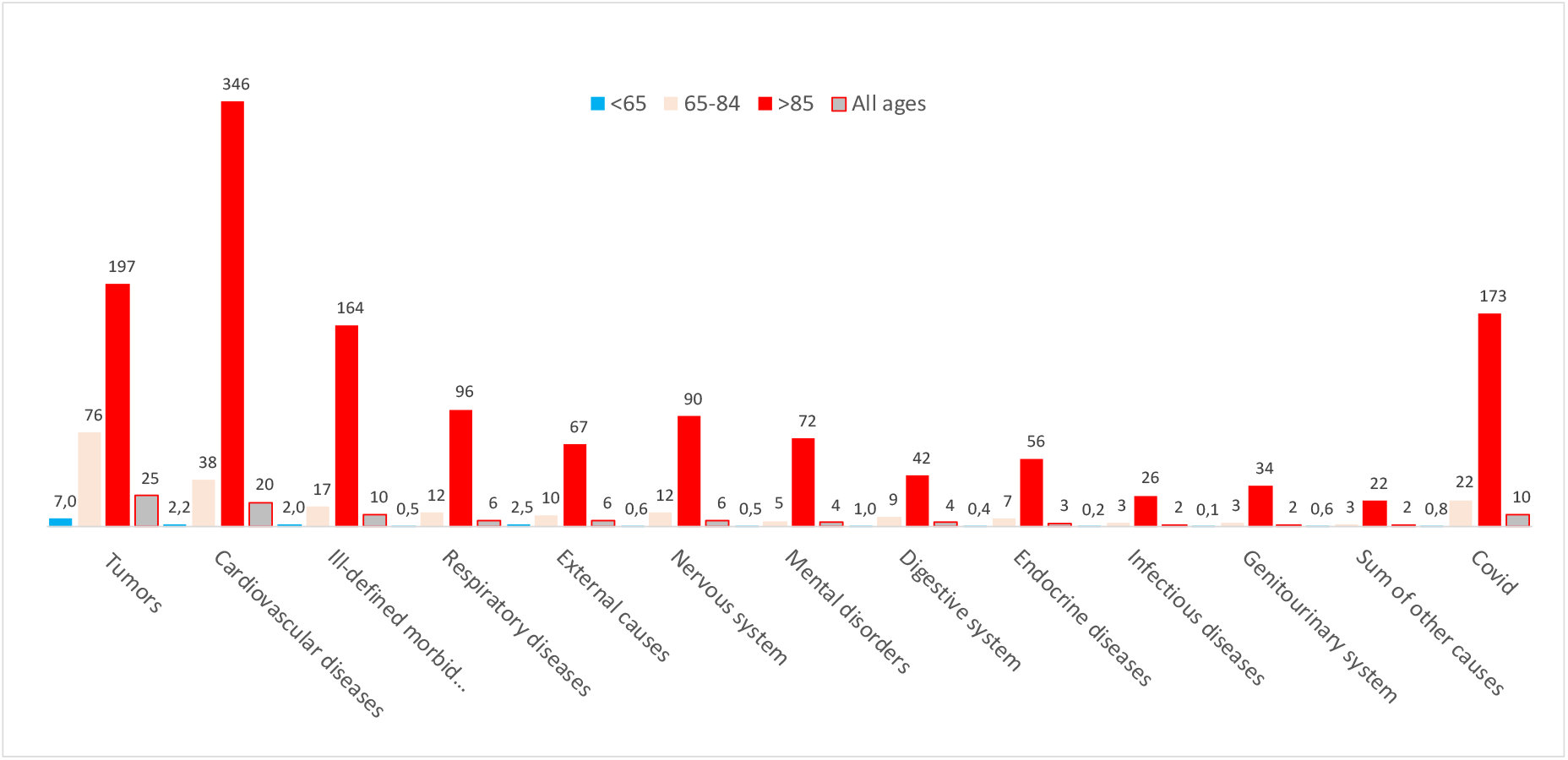
Number of deaths by cause and age group per 10,000 class population in 2020. Source: CépiDc, Insee Demographic Indicators, authors’ calculations. Field: Metropolitan France and its overseas departments and territories Reading note: For each major cause category (on the x-axis), the death rates per 10,000 population of each study class were calculated. For each cause, four bars represent these rates, each with a specific color code: 1) “younger than 65 years old” (in blue), 2) aged “between 65 and 84 years old” (in pink) and 3) “older than 85 years old” (in red) and for the whole population (in gray). For example, the cause “cardiovascular disease” peaks at 346 deaths per 10,000 population over 85 years of age (in red), whereas it is 2.2 deaths per 10,000 population under 65 years of age (in blue).

In France, the results pertaining to the cause “respiratory diseases” are consistent with the analysis of morbidity data, which shows a decrease in the number of hospital stays for respiratory and infectious causes in 2020^29^.

The use of the “Covid-19” label began in hospitals, following instructions from the *Agence Technique de l’Information sur l’Hospitalisation* (ATIH) to report all patients as “Covid-19, virus identified”, even when no test had been performed on the patient10. The hospitalizations recorded as Covid-19 in the “Respiratory infections and inflammations” homogeneous patient groups (GHM) (see ScanSanté^30^) are a simple transfer of hospitalizations previously recorded for other diseases (notably bronchitis, bronchiolitis, bronchopneumopathy, and influenza).

#### Another French exception

France is once again an exception in the study of cause-specific mortality. This may be owing to its historical culture of statistics. In the international literature, analyses of mortality by cause of death in 2020 remain limited. However, the observations show differences between countries^31,32,33^. Unlike France, most countries report an increase in mortality for diseases of the circulatory system in 2020. We have shown that in France, the cause “tumor” decreased significantly regardless of age group. This is far from being the case in the United Kingdom, which reports an increase in tumor^34^ while other countries, such as Italy, the United States, and Mexico, report only a moderate reduction in mortality due to tumors^35,36^. Medical explanations for such disparities are weak. On the other hand, it seems quite clear that labeling bias is key to understanding that the impact of the Covid-19 epidemic was small on the health of the population but particularly significant in France when it comes to the implementation of administrative measures.

## Conclusion

Data on causes of death in France from Inserm’s CépiDc for the year 2020 were made public at the end of 2022. The analysis of these new data is perfectly consistent with the studies conducted based on the INSEE data, the results of which were released at the beginning of 2021. The cause-of-death data thus confirm the minor impact of the Covid-19 epidemic.

The first paradox with “Covid-19” labeled deaths is that the number of deaths labeled with this cause (i.e., 69,000) exceeds the 47,000 excess deaths that were expected. Where did the 22,000 missing deaths come from?

We have shown that they come from other causes which, against all expectations, decreased at the same time, in an abnormal way.

Certainly, questions were raised early on regarding the attribution of the deaths to the epidemic: “Did the individuals die following a severe form of the disease (people who died of the coronavirus infection) or did they die following another disease (people who died with the coronavirus infection)? This distinction is of the utmost importance because the mortality linked to the epidemic is necessarily overestimated if all the deaths of individuals simply suspected of having been infected by the virus are automatically attributed to it. Rather than a major impact of the epidemic, this suggests a competition between the cause “covid-19” and the other major causes of deaths (e.g., tumors, circulatory system diseases). This is precisely what is verified through the analysis of the CépiDc data. It is the extraordinary fact of the study of causes of death in 2020; most of the other major causes of death are significantly decreasing compared to the trends observed over the course of the previous years.

Furthermore, this cause of death affects mainly elderly people. We emphasized that those aged under 65 years have not experienced any excess mortality as it has been the case for more than 20 years except during the H1N1 influenza which, ten years earlier, in a matter of weeks, proved more harmful than the Covid epidemic for this age group. For the older age groups, there is certainly an excess mortality in 2020. This excess mortality has been a recurring fact for more than ten years since the number of elderly people is constantly increasing. But it is lower than the one caused by the 2003 heat wave which, seventeen years earlier, in a matter of days, proved more harmful than the Covid epidemic in one year for these age groups. For all these reasons, the coronavirus epidemic will simply need to be added to the list of the numerous viroses^37^ responsible for severe respiratory pathologies^38,39^. It does not present any major difference with the most severe episodes of seasonal influenza.

However, during this period, a state of emergency was quickly declared. Two lockdowns (i.e., leaving one’s home without permission was prohibited), each longer than 2 months, were enforced. Schools, universities, and most businesses were closed. Curfew periods with traffic restrictions followed. Long periods of mandatory masking in public spaces and other social distancing measures with massive virologic testing campaigns and quarantine were imposed. Numerous studies now show that all these measures were ineffective^40^. The reality is that these measures have been detrimental to all sectors of our society, both for people’s health and the economy of the country.

2020 was marked by the Covid-19 epidemic, which stands out as an unprecedented societal event. The analysis of causes of death in 2020 in France reveals the biases pertaining to the interpretation of “Covid-19” as a cause of death. It makes it possible to understand how administrative rules, implemented during an emergency, have modified the statistics. This analysis confirms the minor impact of the epidemic on mortality. However, the predicted catastrophic increase in mortality was put forward as the main argument for the population to accept extraordinary measures that never in the history of mankind had been implemented on such a scale.

## Data Availability

All data produced in the present study are available upon reasonable request to the authors
All data produced in the present work are contained in the manuscript
All data produced are available online at

https://opendata-cepidc.inserm.fr

https://www.insee.fr/fr/outil-interactif/5014911/pyramide.htm

https://drees.solidarites-sante.gouv.fr/sites/default/files/2022-12/ER1250.xlsx

